# “If we manage early, we can get it right:” Healthcare Workers’ Experiences Managing Sepsis at a Kenyan Referral Hospital

**DOI:** 10.1101/2025.01.23.25321025

**Authors:** Maria Srour, Shamim Ali, Matthew Hodge, Charles Kwobah, Megan S. McHenry, Mary Ann Etling, Amira Nafiseh, Clare C. Prohaska, Babar Khan, Neelima Navuluri

## Abstract

**Background and objectives:** Sepsis and septic shock are conditions of high mortality across the globe. Despite the efforts of the Surviving Sepsis Campaign, improvements in outcomes for patients with sepsis and septic shock have been mostly seen in high-income countries (HICs), as these guidelines are often irrelevant, and in some cases can be harmful in low-resource settings. Thus, low- and middle-income countries (LMICs), still bear most of the global sepsis burden. While this inequity stems from a lack of data from LMICs to inform these international standards, most registered sepsis trials still take place in HICs.

This paper utilizes a socio-ecological model to describe the lived experiences of local healthcare workers treating sepsis and septic shock at a large referral hospital in western Kenya. These perspectives shed light on barriers and strengths in care, gaps in knowledge, and areas of high-yield improvement. This approach allowed us to find potential changes to be made to improve care and patient outcomes.

**Materials and methods:** This is a descriptive analysis focused on providers caring for patients with sepsis and septic shock. Twenty-seven interviews with a wide variety of purposively sampled patient-facing and ancillary medical staff were performed. Concurrent thematic analysis took place as interviews were being conducted. The concept presented were inductively and deductively reasoned and analyzed using a socio-ecological framework. We chose to present three levels of influence on the individual provider.

**Results:** We present our results using a socio-ecological model. At the health system level, we found that most patients to do not have healthcare coverage, which drives up out-of-pocket expenses for individuals. At the hospital level, capacity limits, particularly personnel shortages and small ICU spaces, and influence care. At the interdisciplinary level, relationships between providers and other members of the healthcare team can present challenges. Lastly, these systems-, hospital-, and interdisciplinary-level challenges make guideline adherence difficult and not always feasible for individual providers.

**Conclusions:** To our knowledge, this is the first study to give voice to local providers treating patients with sepsis at a referral center in western Kenya. By presenting findings in the socio-ecological model, we are able to organize potential interventions for the improvement of care at various levels. We found high-yield areas for improving care include establishing clear protocols for task assignments and communication, increasing the number of trained personnel both in the general wards and in the ICU, and on a broader scale, advocating for expanded healthcare coverage for all Kenyans. This work provides a framework for further investigation into elements of sepsis care and the creation of locally relevant treatment guidelines in SSA and across LMICs.

## INTRODUCTION

Sepsis and septic shock are high morbidity and mortality conditions which disproportionately affect individuals in low- and middle-income countries (LMICs) [1,2]. An estimated two million people die every year from sepsis in Africa alone, a number thought to be an underestimate [3]. Since 1991, the global effort to improve the care of adult patients with sepsis and septic shock has centered around the Surviving Sepsis Campaign Guidelines, which were jointly developed by the Society of Critical Care Medicine and the European Society of Intensive Care Medicine. While high-income countries (HICs) have seen improvements in morbidity and mortality, a growing body of evidence shows that implementing the Surviving Sepsis Guidelines in LMICs can be ineffective at best and in some cases harmful [4]. Multiple organizations, including the World Health Organization (WHO) and African Sepsis Alliance, have issued calls for action in decreasing the disparities in sepsis outcomes worldwide. Yet despite this prioritization, very little sepsis-related research data are being generated in LMICs. As of 2018, 55% of registered clinical trials in sepsis were to take place in HICs, despite 85% of sepsis cases and deaths occurring in LMICs, predominantly in sub-Saharan Africa (SSA) and South-East Asia [1,2,5].

This striking paucity of data on sepsis in LMICs creates a challenge in developing treatment guidelines with a broader application. While some principles, such as early and appropriate antibiotics, are generalizable across settings, treatments like intravenous fluid are still controversial and not uniformly beneficial [4,6,7]. Existing literature illustrates considerable heterogeneity in patient populations, illness patterns, and resource-limitations throughout and within LMICs, highlighting the need for context-specific guidelines that are medically appropriate and feasible to implement in variable settings [8,9].

Existing literature on sepsis in adults living in LMICs focuses on epidemiological factors without including the critical voice of local practitioners. These perspectives are essential in understanding the scope of the issues contributing to sepsis care within these settings. This paper utilizes a socio-ecological model to describe healthcare workers’ (HCWs) lived experiences treating patients with sepsis at a large referral hospital in western Kenya. These perspectives could allow us to gain insights into barriers and strengths in care, gaps in knowledge, and potential areas for high-yield improvements. By using this approach, we can elucidate potential changes to be made within current systems to improve care and patient outcomes. To our knowledge, this is the first descriptive study examining the current standard of care for adult patients with sepsis and septic shock at a large referral center in SSA using a socio-ecological model.

## METHODS

### Setting

This study took place at Moi Teaching and Referral Hospital (MTRH), a 900-bed hospital with a catchment area of nearly 25 million people, in Eldoret, Kenya. The hospital includes three Intensive Care Units (ICUs): one larger medical and surgical unit with approximately 25 beds, one Cardiac Care Unit (CCU) with approximately 12 beds where patients can receive vasopressors but cannot be invasively mechanically ventilated, and a smaller six bed medical ICU Given the hospital’s status as a national referral center and its partnership with the Academic Model Providing Access to Healthcare (AMPATH), a consortium of 14 North American academic institutions, along with MTRH, and the Moi University School of Medicine, it is a relatively well-resourced hospital within Kenya’s public sector [10]. As a public hospital, some services are free, however patients are still required to financially cover the majority of their care costs. As of 2018, 19.9% of Kenyans had some form of health insurance. The National Health Insurance Fund (NHIF), a governmental organization whose “core mandate is to provide medical insurance cover to all its members,” covered 94% of those who were insured [11]. While NHIF was in existence during the study period, Kenya transitioned from NHIF to the Social Health Insurance Fund (SHIF) in 2024 [12].

### Design

This is a descriptive study which used semi-structured interviews to assess the experiences and perspectives of healthcare workers caring for patients older than 14 years with newly suspected or diagnosed sepsis and septic shock on the General Internal Medicine (GIM) wards at MTRH. Data was collected and analyzed using a socio-ecological framework.

### Recruitment and data collection

Medical teams at MTRH often consist of fourth- and sixth-year medical students, medical officers, clinical officers, nurses, pharmacists, Internal Medicine registrars, and consultant physicians (Table 1). All provider types caring for patients who were both admitted through Casualty (Emergency Department) or as transfers from outside hospitals, as well as phlebotomists and microbiology laboratory technicians were eligible for participation. Participants were recruited using purposive sampling of all patient-facing members of the GIM teams on the wards at the conclusion of morning rounds or as pre-arranged meetings [13]. Participants were informed about the goals of the study prior to the interviews. Trainees were reassured that this was not a graded exercise and would not influence any academic scores. All questions and concerns from participants were addressed and written informed consent was obtained.

**Table 1:** Qualifications of Healthcare Workers in Kenya.

| **Role** | **Qualification** |
| --- | --- |
| Clinical officers (COs) | Clinical Officers are required to complete a three-year professional diploma. They are licensed by the Clinical Officers Council have unlimited practice rights in Kenya. |
| Consultants | Consultants have completed the Bachelor of Medicine and Bachelor of Surgery (MBChB) degree as well as a master's in medicine in Internal Medicine and act as supervising physicians. |
| Internal Medicine (IM) registrars | Registrars have completed the MBChB degree and are in postgraduate training to earn a master's in medicine in Internal Medicine degree through Moi University. |
| Medical officers (MOs) | Medical Officers have completed the MBChB degree with no postgraduate clinical training. |
| Medical student | Students enter a six-year MBChB program after completion of high school. Clinical rotations occur in years four through six. |
| Microbiology Laboratory Technicians | Laboratory technicians earn a certificate after two years of training at one of 25 institutions approved by the Kenya Medical Laboratory Technicians and Technologist Board. |
| Nurses | The Nursing Council of Kenya recognizes four categories of nurses with varying qualifications. Nurses can have a certificate, diploma, Bachelor of Science in Nursing degree, or a Bachelor of Science in Midwifery degree. All must be licensed by the Nursing Council at their respective levels. |
| Pharmacists | Pharmacists at MTRH have completed a five-year didactic Bachelor in Pharmacy program. Some are completing a three-year master's in pharmacy degree through Moi University. |
| Phlebotomists | Phlebotomy courses are offered throughout the country and range in duration from several days to months. |

Interviews were conducted by the lead investigator (MS) or one of two research assistants (RAs). Interviewers used one of two standard interview guides for either patient-facing providers or ancillary workers (see appendix). Both guides were developed with the help of Kenyan GIM consulting physicians at MTRH serving as co-investigators for the study. At the time of study, the lead investigator was in her final year of Pulmonary and Critical Care Medicine (PCCM) fellowship at an institution in the United States and had previously rotated at MTRH as a student and trainee, with appropriate mentorship. Both RAs were U.S. MD/MPH students with specific qualitative methods training. Due to space and time constraints, interviews for all non-consultant providers took place in quiet areas near the GIM wards. Consultants, phlebotomists, and microbiology laboratory technicians were interviewed in private offices.

Interviews were performed in English and audio recorded. English is a national language of Kenya and all healthcare workers (HCWs) at MTRH are fluent. Transcripts were generated and edited by MS for accuracy. Field notes were also taken during the interviews. Average interview time was 14:27 minutes. Interviewers did not repeat interviews, share transcripts with participants or approach participants for feedback regarding their responses. Interviews were completed between January and May 2023.

### Data management and analysis

The study team, including MS and MH, an Internal Medicine Chief Resident at an American institution who had completed clinical rotations at MTRH, analyzed interview data using the socio-ecological model. The socio-ecological model recognizes the interplay between environmental, social, and communal factors and individual human behavior [14]. Such a model allows for categorization of interventions that can be reasonably addressed using quality improvement methods, and other issues that must be addressed with policy and higher-level decisions. For this analysis, the socio-ecological model was modified to include three levels of influence on the individual provider. Most immediate to the provider is the interdisciplinary level, then the hospital, and finally the Kenyan healthcare system.

Interview transcripts and field notes were reviewed by MS and MH, who then used NVivo 13 to independently complete line-by-line intermediate selective coding, using the codebook generated by MS [13]. They then came together to discuss key areas of congruency and agreement until consensus was reached.

### Research ethics approvals

All procedures and protocols were reviewed and approved by the MTRH/Moi University Institutional Research Ethics Committee as well as the Indiana University Institutional Review Board. This study did not receive monetary funding support.

## RESULTS

Twenty-seven total interviews were conducted with a clinical officer (n=1), consultants (n=4), Internal Medicine registrars (n=7), medical officer (n=1), medical student (n=1), microbiology laboratory technicians (n=3), nurses (n=4), pharmacists (n=3), and phlebotomists (n=3). Fifty-four percent of the participants who were asked self-reported as male, 46% female. Mean time in position ranged from 8 months for COs to 12 years for nurses. These data are outlined in Table 2. Our description of HCWs’ experiences is presented through the framework of the modified socio-ecological model as outlined in Figure 1.

**Table 2:** Participant Characteristics.

| Position | Number of Interviewees | Self-reported Sex |  | Mean Time in Position |
| --- | --- | --- | --- | --- |
|  |  | Male | Female |  |
| Clinical Officers | 1 | 0 | 1 | 8 months |
| Consultants | 4 | 3 | 1 | 8 years |
| Internal Medicine Registrars | 7 | 4 | 3 | 2 years |
| Medical Officers | 1 | 1 | 0 | 3 years |
| Medical Students | 1 | 1 | 0 | 6 years |
| Microbiology Laboratory Technicians | 3 | 1 | 2 | Not collected |
| Nurses | 4 | 0 | 4 | 12 years |
| Pharmacists | 3 | 3 | 0 | 10 years |
| Phlebotomists | 3 | Not collected | Not collected | Not collected |
| **Total** | **27** | **13** | **11** |  |

### Health System Level: A lack of insurance coverage increases patients’ out-of-pocket expense

Cost to the patient affects how HCWs approach the workup of sepsis, initial management, and escalation of care. Due to cost restrictions, workups tend to be more judicious, treatment choices are reflective of cost, and private ICU transfers are often prohibitive.

#### Workups are slower and more judicious

Patient costs for laboratory or radiographic investigations often depend on the test, and many clinicians work with a basic knowledge of what is covered by the hospital and what must be paid by the patient and their families. As one consultant described, “For the things that are automatically billed on patients’ hospital bills, you would request them without thinking … But then for the ones that sometimes patients have to pay beforehand, those ones have delays.” One registrar described his process when ordering procalcitonin, a biomarker that has been used to shorten antibiotic durations, “When I’m going [through] the history and find that they have insurance then I don’t hesitate doing the procalcitonin [15].”

One consultant described, “Yes, you’ve made a diagnosis or you have a high index of suspicion for sepsis, what you do next is where now we see the gaps … It’s almost done sequentially. Get a chest X-ray. If that’s ok, then now get a urinalysis, and then look for another source. Which sort of creates a delay.” Providers shared that this serial form of diagnostic workup, as opposed to sending multiple investigations at once, is often driven by cost-consciousness and reluctance to add fees for unnecessary tests, especially for indigent patients. These concerns for cost can often dictate clinical decision making and take precedent over guideline-or evidence-based practice.

#### Treatment choices reflect cost

Cost of treatment for patients without insurance coverage can influence medication selection and treatment duration as well. One registrar noted, “Certain antibiotics-although they are available, sometimes they might end up being costly for patients who don’t have NHIF cover.” Another gave a similar example. “We did give the patient the polymyxin B [a relatively expensive antibiotic]. But later on, the patient actually had problems when trying to clear the bill because they don’t have NHIF, because it’s sometimes an expensive drug … So you find that most patients, they’re in the ward, but they keep asking you to discharge them earlier, even before they have actually the sepsis resolved, because they want to reduce the bill.”

Ongoing management is also affected by cost and is generally focused on clinical observation of the patient. For example, laboratory testing is rarely done daily. As a consultant described her practice, “Follow the labs, but more importantly, follow the patient. So let’s say whatever made us think of sepsis-if they turned around fairly quickly, like afebrile in 48 hours, I probably repeat my inflammatory markers then just to show that they’re coming down on a downward trend … I try and use the shortest amount of duration possible for that specific infection.”

#### Private ICU transfer is cost prohibitive

Patients who are critically ill and cannot be admitted to the ICU at MTRH, typically due to lack of beds, are given the option of finding a private facility in town. Cost of this treatment must be assumed by the patient and family. If this is not feasible, they are asked to sign a consent form to continue receiving ward-level care. A nurse described the process, “We advise the relatives to source from the outside if the relatives are able to pay for the services outside the hospital … in case you don’t have an ICU and the patient needs an ICU, there is a consent form. They have to sign that you have been told there is no ICU [space] in our facility and you have to look for an external. But since they don’t have it, they sign the form in case.”

This expensive option is rarely feasible for patients at MTRH. “Patients who are admitted in this ward come from very poor backgrounds. So they can’t even afford outside ICU, so we are managing them in the wards,” another nurse added.

### Hospital Level: Capacity limits influence care

Significant capacity limitations still exist at MTRH. Participants noted major challenges in personnel shortages and restricted ICU space, despite ongoing improvements. These subthemes are closely linked, as personnel shortages have significant downstream effects.

#### Personnel shortages cause treatment delays and limit bed availability

“We just try to advocate for early recognition and management of sepsis … If we manage early, we can get it right,” one nurse said. Given staffing ratios, however, this can be difficult. A consultant explained her workflow stating, “I think the patient numbers are overwhelming. So if you’re in a team and you have 45 patients under your care it’s easy to have some things fall in the gap, especially following up results and stuff.”

Treatment delays are evident. One nurse stated, “This is a public hospital. The staffing ratio in relation to the patient ratio is not adequate … You will concentrate on the patients in the acute room. When you go to the other end, maybe there’s a delay … Say a patient is on IV fluids and is supposed to get four liters. So maybe you’ll have a deficit because of the timing. You are engaged. So you may end up not infusing your patients very well.”

The staffing ratio challenge extends to support staff as well. In phlebotomy, a team of five covers close to 200 medical patients. In general, phlebotomists are responsible for drawing all blood cultures in medical wards and casualty. Beyond treatment, bed availability can also become an issue. In describing the availability of ICU space, one consultant noted, “We don’t have enough nursing staff. So currently we are limiting ourselves.”

#### ICU space is restricted

Though the number of ICU beds at MTRH increased in response to the Covid-19 pandemic, capacity is still quite limited and not enough to provide care for most critically ill patients. As one registrar described, “The problem with that is normally we have limited space in the ICU. So we are just forced to manage them in the wards.”

One consultant who practices in the general medicine wards described the challenge of finding ICU space as such: “A lot of tears and prayers. That’s like first entry criteria. I think the surgical departments and obstetrics and pediatrics … it almost sometimes feels like there’s preferences to admit from those departments than from medical ward … Because of our limited ICU resource, it ends up being like weighing which patients will benefit more and unfortunately with our population, we don’t seem to fit that bill.” A nurse describes, “If you want to get an ICU space there, they either need to discharge a patient or a patient has to succumb … If there is a space, then your patient will be lucky.”

### Interdisciplinary Level: Strong multidisciplinary relationships impact care

Streamlined processes that incorporate multidisciplinary personnel help in assigning specific roles and eliminating gaps in care. Respondents stressed the need for clearer communication protocols and an emphasis on teamwork.

#### Teamwork is important

One pharmacist described his experiences working with physicians on the wards saying, “Our medical doctors are very open. They are not rigid to any regimen … If they don’t agree then they would differ politely. It’s not like our ideas are being thrown to the dust.” As one registrar noted, “there is teamwork in the wards.”

Still, some issues of hierarchy still arise. A consultant cited fear of escalating care for ICU transfer as a possible point of contention. “It is usually an anesthetist or a medical officer. We have one respiratory therapist [in ICU] … So you can imagine we have a challenge there. I’m a physician. I feel I’m okay. So sometimes I don’t feel okay asking somebody to come and agree with me.”

A nurse echoed the other side of this issue. “That fear of speaking … But that’s people who don’t know how to advocate for patients. There are some who have fear because people are above us and sometimes you think, ‘How can I speak out?’ But we [nurses] should.” A medical student summed up the need for interdisciplinary work. “Develop a concrete protocol for treating sepsis-one that incorporates everyone so that everyone knows their role. Doctors, residents, students, nurses, everyone has to know their role.”

#### Communication with the microbiology lab is a pressure point

Timely result reporting from the microbiology lab was another widely recognized issue. “We do have a problem. Honestly, we do have a problem with cultures. Because ideally three days you should already have the cultures or at least a gram stain. But we normally get them after five days, seven days, even up to two weeks,” one registrar said.

Some respondents reported that the job of communicating culture results belongs to the phlebotomists who physically carry samples from the wards to the microbiology lab. As one phlebotomist described, “Our results are still paper-based … So normally we have to pick the physical result and bring it to the team that’s working, so we literally just hand over to clinicians. We don’t really communicate the results. We just hand over two papers.”

This practice is not standardized. As a microbiology lab technician described, “That scenario is somehow very difficult to handle because it is something that you cannot say that [a] specific person is responsible for this kind of communication.” A nurse echoed this description when discussing result reporting saying, “It is not consistent.”

One consultant commented with possibilities for change. “If the lab could communicate … When you have abnormal electrolytes, they call and say we’ve flagged a hypokalemia of 2.3. If we grew an organism, I think they could maybe alert the ward. Or maybe we could have a policy such that every morning all the results from culture come.” Another nurse summed up the issue stating that “If communication, like from the lab and the nursing and the doctors can be improved, it can improve the outcome of our patients too. Communication, communication. That’s the biggest thing.”

### Individual Provider level: Guideline adherence is not always feasible

While many providers were aware of recommendations made by the Surviving Sepsis Campaign and other guidelines, clinical practice seemed to be driven by local culture and feasibility. Hence, protocols for sepsis workup are not standardized and treatment patterns vary widely.

#### Workup protocols are not standardized

Several trends were noted in the general practice culture for the workup of sepsis. Most clinicians begin with presenting symptoms and physical exam. “You check for signs of sepsis like fever. Systemically you’ll have tachycardia and hypotension,” said one registrar. Basic workup is generally similar across providers and includes “the usual-the blood counts, the kidney function,” as described by one consultant. One seemingly habitual practice is that of checking C-reactive protein (CRP) and procalcitonin. One registrar cited a situation in which CRP was used diagnostically early on. “The only thing that made us think about sepsis was that we had some high CRP, and we couldn’t get any other any other reason for his altered mental state. He’s been on dialysis; he has a dialysis catheter. We thought that [central line-associated bloodstream infection] could be it, most likely … We gave him some antibiotics and he got better.” A consultant described his use of inflammatory markers for trending patient progress. “Most of the time, I use it early on to figure out whether we have a bacterial infection or not. Once we feel like we’ve done an adequate duration of antibiotics, we check the CRP and the procalcitonin again to just see it coming down. And then that gives us a sense of when we need to stop antibiotics.”

Serum lactate is not followed routinely. One registrar describes, “the trouble we have with lactate is that you only get it on a blood gas. We don’t have a lactate measured by the lab as another thing. You need to do a blood gas.” If the patient is not in respiratory distress, the other parameters measured on the blood gas analysis are unlikely to change management and are thus seen as wasteful. As another registrar explains, “Honestly, I really don’t order for lactic [acid] because we can only get it from the blood gas. So if they don’t come with resp distress, hypoxia then we’re not going to be able to get our blood gas. But ideally, we should be doing that.” Laboratory technicians further clarified that MTRH does have the capability to check serum lactate in the chemistry lab without a blood gas analysis, but the reagent is often left unused until the expiration date as few clinicians are aware that it is available.

Collecting early blood cultures is a practice many noted has improved in recent years, but no strict hospital policy exists. As one consultant noted, “it would be good if we had a policy whereby if the clinician handling the patient at the first point of contact suspects that this is an infectious process, then you should take samples [for blood cultures] at that point, before you start any antibiotics.”

Another consultant described the process of further workup beyond basic sepsis recognition. “I think the gaps appear in terms of now the next step. Yes, you’ve made a diagnosis, or you have a high index of suspicion for sepsis. What you do next is where now we see the gaps in terms of getting the appropriate labs.”

#### Treatment patterns vary by provider

Treatment practices were noted to be heterogenous across the medicine wards. “I think you have the individualized approach to treatment because it literally depends on who you’ve learned under and who you’ve worked with,” one consultant noted.

Most participants reported antibiotics as the mainstay of treatment. However, choosing an appropriate and effective empiric regimen after taking the patient’s history into account can be challenging in this setting. As one pharmacist explains, “Once the patient lands on the outpatient setup, before they get admitted, they are always given antibiotics. By the time they land on the ward, these patients are on antibiotics already … If they’re coming from another hospital then we would first think that this patient already has received ceftriaxone from the community, or even amikacin from the community or gentamicin from the community. So we are likely to shift to a second line which is now cefepime.”

One consultant described her attempt to remain judicious in antibiotic choices. “I’ve been trying to protect the carbapenems as much as possible, but I think it’s almost a losing battle because I think everyone else is like, ‘You have a fever, you’ve been in the hospital, here is some meropenem for you!’”

Improvements have been noticed lately. Multiple providers including consultants and registrars commented on a new antibiogram being developed for the hospital. A few expressed hopes that this would ease the difficulty in choosing empiric antibiotics for their patients.

Intravenous fluid resuscitation did not seem to have a routine practice. Some providers cited three liters in 24 hours, while others noted that they simply watch blood pressures. One registrar commented, “If they come in septic shock, we usually try to get 30ml/kg like what is recommended, but you rarely get that much fluid given.” Ultimately, treatment practices were found to be heterogenous across the medicine wards. “I think you have the individualized approach to treatment because it literally depends on who you have learned under and who you have worked with,” one consultant noted.

## DISCUSSION

This descriptive study presents several barriers and facilitators in caring for adult patients with sepsis and septic shock through the perspectives of local HCWs. Our results identify areas for intervention and quality improvement at the health system, hospital, interpersonal, and individual provider levels. At the health system and hospital levels, alleviating out of pocket expenses for patients and increasing the pool of trained personnel can improve care. Interdisciplinary and individual provider interventions, such as streamlining communication protocols and emphasis more efficient equipment storage, could be more immediate solutions.

At the health system level, cost to the patient significantly influences care. In line with the United Nations Sustainable Development Goals, Kenya has committed to providing its citizens with universal health coverage by 2030 [16]. Still, NHIF is used by fewer than 20% of Kenyans, payouts provided by NHIF are limited and far from comprehensive, and close to 20% of Kenyans live more than one hour away from an NHIF-contracted inpatient facility [11,17,18]. This lack of coverage, even at a public facility, creates significant challenges and delay in care at tertiary facilities where providers are conscientious of patient fees. In our study, many clinicians also reported sending broader workups when a patient was known to have NHIF coverage. Our data show insufficient healthcare coverage leads to more judicious care, which can lead to life-threatening delays in acute cases such as septic shock. In October 2024, Kenya began transitioning from NHIF to the Social Health Insurance Fund (SHIF). So far, the transition have been chaotic and disorganized, creating more delays and prohibitive costs for patients seeking care [12]. While the overall success of SHIF remains to be seen, a future in which more citizens have access to affordable care and are knowledgeable about their coverage options is imperative to delivering timely services [19].

At the hospital level, the most pressing issues are personnel shortages and ICU space. While the number of healthcare workers in Kenya increased by 110% from 2010 to 2020, this has not resolved the dire need for more trained professionals, and such advances are often overshadowed by emigration of young professionals in Kenya’s long-established brain drain [20,21]. While we echo calls to increase training opportunities for all healthcare workers in Kenya, there is a need for more short-term solutions to the larger personnel problem.

Shifting non-medical tasks away from physicians and nurses, streamlining care and interpersonal interactions, and optimizing hospital and equipment layouts can help to improve the available care in the meantime [22,23]. Furthermore, the results of our study complement quantitative studies reporting on the availability of ICU resources in Kenya [24,25]. The paucity of ICU resources reflects not only a lack of physical space and equipment, but more importantly, a lack of personnel with critical care training [26]. As of 2023, the Emergency Medicine and Critical Care Clinical Officer training program established in 2017 at Kijabe Hospital, a 350-bed faith-based tertiary care hospital in central Kenya, has graduated 35 clinicians since its inception in 2014. This program has been replicated at the Kenyan Medical Training College and now has a pediatric equivalent [27]. Continuing to replicate similar programs, making them accessible through public hospitals, and expanding training to physicians, nurses, and ancillary ICU personnel such as respiratory therapists, is a crucial step in improving Kenya’s critical care capacity.

Regarding interdisciplinary relationships, clear communication protocols can alleviate several of the issues identified at the interdisciplinary level. For example, our data show that the role of communicating laboratory results is not assigned to one person, nor is there an established expectation for the timing of reporting these results. Timely reporting of laboratory results has been noted as a high priority area in building laboratory capacity in LMICs [28]. Implementation of established quality improvement methods, such as assigning clear tasks and expectations, and even employing secure text messaging, could improve communication between clinicians and ancillary staff, with the potential of providing efficient care, and ultimately decreasing mortality [29,30].

Finally, the health systems and hospital level barriers pose serious challenges to individual providers on the medical wards. The result is a heterogeneous treatment pattern across the wards, highly dependent on the clinician providing care for the patient. Standardized sepsis protocols in resource-constrained settings are controversial but illustrate that realistic guidelines that do not add to workloads are crucial [9]. Simple measures such as keeping vitals machines on the wards, accessible storage of IV tubing, blood culture bottles, and commonly used empiric antibiotics in patient care areas, ensuring that blood culture bottles are sufficiently stocked, and informing clinicians about what test reagents and medications are available each day may streamline care, allowing for timely treatment after recognition.

This study has several strengths. Interviews were done with a wide range of HCWs at MTRH, allowing us to describe the current standard of care from multiple perspectives. Interviews with patient-facing providers were semi-structured and conducted by a single investigator who continually reviewed the data. This method allowed for earlier and deeper probing into specific issues that emerged as significant themes early on. Use of the socio-ecological model allowed us to identify barriers, strengths and knowledge gaps on several levels to provide a system-wide perspective and description. Some limitations exist. First, due to the time constraints and clinical demands of some participants, our interviews were brief which may not have allowed for the in-depth probing possible in a longer interview. Second, because of the demands of clinical care, despite efforts to use private spaces, many interviews were conducted in quieter spaces on the wards without complete privacy. However, interviewing providers in their space of work provided for more widespread participation across HCWs. Third, the interviews and analysis were completed by North American investigators. To mitigate bias, Kenyan co-authors helped to design and implement the study, reviewed the final results, and agreed that our analysis depicts a realistic description of provider experience. Finally, this is a single-center study that focuses on practices in one medical ward in a large referral hospital in a middle-income country, which limits its generalizability, though existing literature shows many similar challenges across similar settings.

## CONCLUSION

Our study amplifies the perspectives of local providers treating patients with sepsis at a national referral hospital in western Kenya and highlighting challenges at the health systems, hospital, interdisciplinary, and individual provider levels using the socio-ecological model. We found high-yield areas with the potential to improve care including establishing clear protocols for task assignments and communication, increasing the number of trained personnel both on general wards and in the ICU, and on a broader scale, advocating for expanded healthcare coverage for all Kenyans. This work provides a framework for further investigation into elements of sepsis care and the creation of locally relevant treatment guidelines in SSA and across LMICs.

## Data Availability

All data produced in the present study are available upon reasonable request to the authors.

---

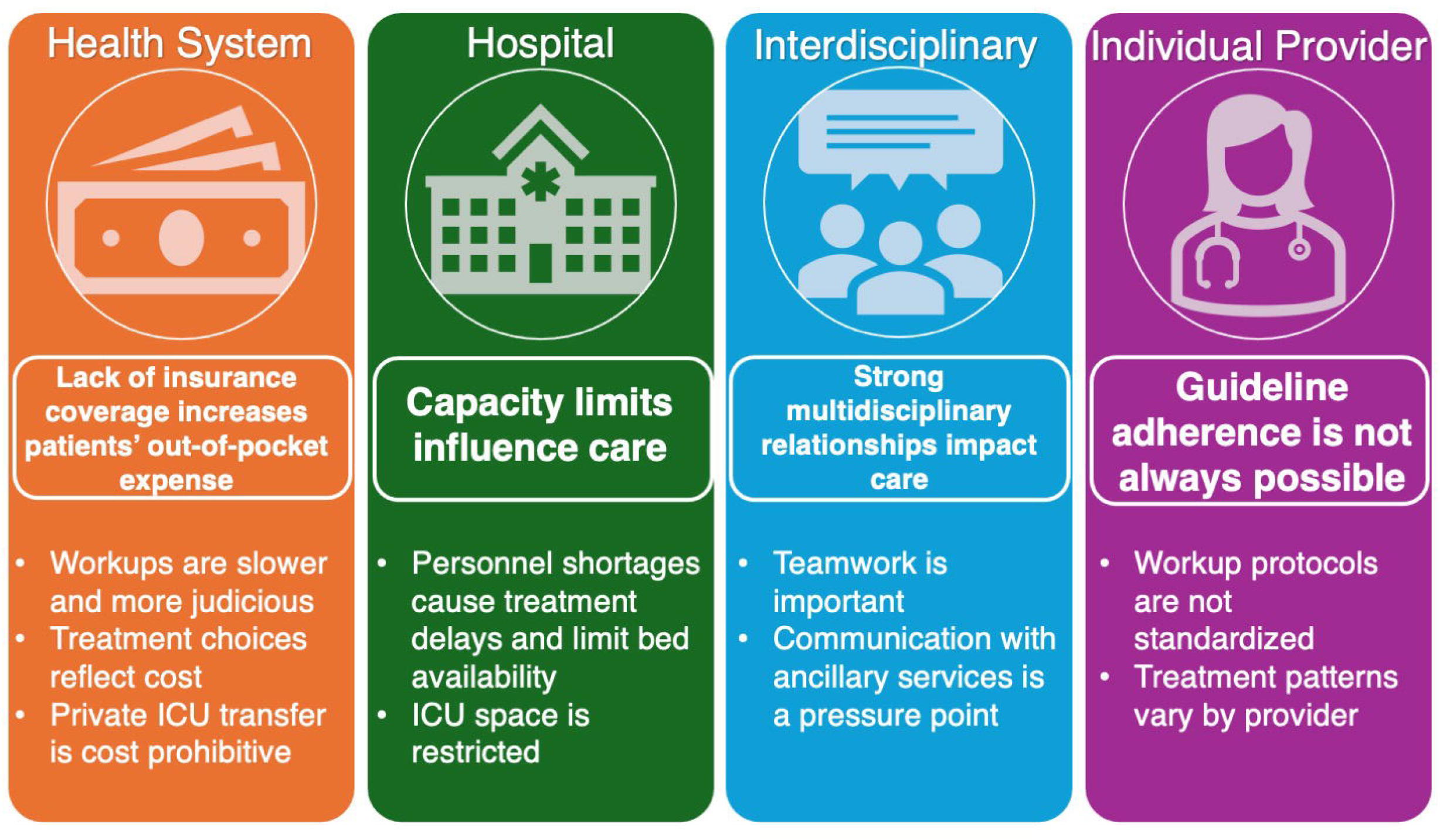

